# Global Assessment of IRF8 as a Novel Cancer Biomarker

**DOI:** 10.1101/2021.12.06.21267361

**Authors:** Daniel C. McQuaid, Gauri Panse, Wei-Lien Wang, Samuel G. Katz, Mina L. Xu

## Abstract

Interferon regulatory factor 8 (IRF8) is a member of the IRF family that is specific to the hematopoietic cell and is involved in regulating the development of human monocytic and dendritic-lineage cells, as well as B cells. Since its utility as a sensitive and specific monoblast marker in the context of acute monocytic leukemias has been recently demonstrated, we hypothesized that it may also be useful as a novel immunohistochemical marker in myeloid sarcomas and blastic plasmacytoid dendritic cell neoplasms (BPDCN) with respect to their differential diagnoses. In this retrospective study, we analyzed the IHC expression pattern of IRF8 in 385 patient samples across 30 types of cancers, referenced to their mRNA expression data available through TCGA. In addition, we assessed IRF8 in 35 myeloid sarcomas, and 13 BPDCNs. Twenty-four of 35 cases of myeloid sarcomas (68.5%) showed positivity for IRF8, with six cases (17.1%) demonstrating IRF8 expression in the absence of CD34 and MPO. All 13 of 13 BPDCNs (100%) showed strong uniform expression of IRF8 and was occasionally more definitive than CD123. IRF8 was negative in all desmoplastic small round cell tumors, Ewing sarcomas, synovial sarcomas, and undifferentiated pleomorphic sarcomas, as well as all epithelial malignancies tested except for 2 triple negative breast cancers that showed subset weak staining. In conclusion, IRF8 is a novel marker helpful in identifying extranodal hematopoietic tumors that can otherwise be difficult to diagnose given the broad differential diagnoses and frequent loss of more common lineage-defining markers.

## 1.0: Introduction

Interferon regulatory factor 8 (IRF8) is a hematopoietic-specific member of the IRF family that is essential in the commitment of myeloid progenitors to monocyte, macrophage, and dendritic cell lineages (1, 2). IRF8 expression is significantly elevated in monocyte-dendritic cell progenitors in mice and conversely, *Irf8*^-/-^ mice lack bone marrow resident macrophages, dendritic cells, and plasmacytoid dendritic cells in lymphatic tissue (3-5). The role of IRF8 in the generation of monocyte and dendritic cell progenitors is conserved in humans, as mutations that impair IRF8 transcriptional activity are associated with immunodeficiency via decreased monocyte and dendritic cell populations (6). Past work has shown that IRF8 is upregulated in subsets of acute myeloid leukemia (AML) and correlates with poor prognosis in AML patients (7). Moreover, we recently demonstrated that IRF8 immunohistochemistry stains monoblasts in cases of acute monocytic leukemia and advanced chronic myelomonocytic leukemia, but is negative in mature monocytes and granulocytes (8). Given this observation, IRF8 may hold promise as a biomarker for additional malignancies derived from early monocyte and dendritic cell precursors.

Myeloid sarcoma is a less common hematologic malignancy of immature monocyte and/or granulocyte populations that manifests as an extramedullary soft tissue mass (9). It is defined by the World Health Organization as a tumor of myeloid blasts that occurs at an anatomic location beyond the bone marrow (10). Myeloid sarcoma typically develops secondary to AML, although rare cases of isolated disease without bone marrow involvement or history of myeloid malignancy have been described (11). These tumors most commonly arise at the lymph nodes, skin, soft tissues, and bone, and must cause effacement of local tissue in order to render a myeloid sarcoma diagnosis (10). Due to the disparate sites of disease involvement, clinical presentation is varied and pathology evaluation is critical for diagnosis. However, myeloid sarcoma is notoriously difficult to diagnose, with an estimated misdiagnosis rate of 25-47% (12-14). This diagnostic inaccuracy is a product of disease rarity, particularly in the absence of a known myeloid disorder, as well as a dearth of markers for staining (15). Histologically, the tumor demonstrates myeloid cell infiltration with myeloblasts or monoblasts (16). Of note, lesions with granulocytic differentiation have features such as eosinophilic metamyelocytes that aid in a myeloid sarcoma diagnosis, while tumors of monocytic origin lack these characteristics and are typically more difficult to diagnose (12). Consequently, additional biomarkers are needed to ameliorate the diagnostic challenges of this extramedullary malignancy.

Blastic plasmacytoid dendritic cell neoplasm (BPDCN) is an aggressive hematopoietic disorder of plasmacytoid dendritic cells (17). BPDCN is a rare cancer, estimated to account for less than 1% of all hematologic malignancies, or roughly 700 cases annually in the United States (18, 19). In 90% of patients, BPDCN presents with skin manifestations and is later accompanied by involvement of bone marrow, peripheral blood, and lymph nodes (20, 21). Progression to systemic disease is rapid, and median survival is estimated to be between 12 and 14 months (22). Diagnosis of BPDCN relies on clinical presentation, histological evaluation of the lesion, and immunophenotyping (23). Due to cutaneous involvement that presents with blast cells in the dermis, suspicion is often high for soft tissue tumors, T-cell leukemias/lymphomas, and NK-cell leukemias/lymphomas (22). It can be difficult to differentiate these malignancies from BPDCN, and immunophenotyping is critical to validate the observed skin lesions are comprised of plasmacytoid dendritic cells rather than myeloid or lymphoid blasts (24). Additionally, expression of antigens expressed by other cell lineages, such as CD56 on NK/T-cell leukemia/lymphomas, and CD4 and CD123 on AMLs can make it difficult to render a definitive diagnosis (25). Thus, additional immunohistochemical markers in the diagnostic panel of BPDCN can help differentiate this malignancy from others with similar morphologic characteristics and clinical presentations.

We hypothesized that IRF8 may serve as a useful biomarker for myeloid sarcoma and BPDCN due to their prominent monoblastic and dendritic cell origin, respectively. Herein, we demonstrate that IRF8 is able to stain neoplastic cells of both myeloid sarcoma and BPDCN, even in cases lacking traditional markers for these malignancies. Importantly, IRF8 expression is absent in a panel of soft-tissue cancers that could potentially mimic myeloid sarcoma and BPDCN, as well as solid tumors. Our work demonstrates the utility for IRF8 in the identification of these rare hematologic malignancies that are in need of additional markers to aid in diagnosis.

## 2.0: Methods

### 2.1: Human Tumor Samples and TMA Creation

35 cases of myeloid sarcoma and 13 cases of BPDCN were obtained following approval by the Yale institutional review board. All cases were diagnosed by board-certified pathologists at the Yale School of Medicine according to their WHO criteria and validated by a second reviewer at outside institutions. Both a pan-cancer tissue microarray of 150 cases spanning 25 tumor subtypes and 27 tissue-matched normal samples as well as a triple-negative breast cancer-specific TMA of 97 cases were made in the Department of Pathology at the Yale School of Medicine. All cases were previously diagnosed at Yale and utility of these biopsies in this study was approved by the Yale institutional review board. In addition, we used specimens from select sections of various previously published sarcoma-specific TMAs, including desmoplastic small round cell tumor (n=30), Ewing sarcoma (n=24), synovial sarcoma (n=45), and undifferentiated pleomorphic sarcoma (n=12).

### 2.2: IHC Staining and Scoring

Immunohistochemistry (IHC) of all aforementioned samples and TMAs was performed using a rabbit monoclonal IRF8 antibody (Anti-IRF8 ab207418 from Abcam, 1:900 dilution) as previously described (8). We additionally stained myeloid sarcoma samples with both myeloperoxidase (MPO rabbit polyclonal concentrate from Dako, 1:10 000 dilution) and CD34 (QBEND from Ventana, Neat), and BPDCN samples with CD123 (9F5 from BD Biosciences, 1:50 dilution). Cases that were double stained with IRF8 and lysozyme utilized the double-staining protocol on Bond with lysozyme (polyclonal from Agilent, Neat). To perform staining, formalin-fixed, paraffin-embedded tumor tissue sections were deparaffinized and rehydrated prior to antigen retrieval and primary antibody incubation. Afterward, specimens were incubated with diaminobenzidine (DAB) chromagen for primary antibody detection and counterstained with hematoxylin. All stained samples were evaluated for both percentage of immunoreactive cells and staining intensity (0, negative; 1+, weak; 2+, moderate; 3+, strong) (26). When possible for both TMAs and whole tissue sections, at least 3 non-adjacent fields containing tumor cells were evaluated for IRF8 staining. Only IRF8 nuclear staining of neoplastic cells was considered positive, as several samples demonstrated cytoplasmic staining of cancerous cells or nuclear staining of infiltrating lymphocytes. Nuclear reactivity at any intensity was denoted as positive for IRF8 and given as percentage of overall tumor cells in a sample. Percentage of positive cells within tumor was also scored for CD34 and MPO in all myeloid sarcoma cases.

### 2.3: TCGA Database Analysis

Expression levels of IRF8 across different cancer types and their matched normal tissue were analyzed using Gene Expression Profiling Interactive Analysis (GEPIA), which utilizes global RNA-Seq data from The Cancer Genome Atlas (TCGA) and Genotype-Tissue Expression (GTEx) databases. While TCGA contains expression data from 33 types of cancer, we collated this dataset to only analyze subtypes that were represented in our pan-cancer and sarcoma TMAs. We then determined the log2 fold change of IRF8 expression between cancerous and normal tissue, utilizing a fold-change threshold of 1 and a p-value cutoff of 0.01 (as determined by a one-way ANOVA) to identify significantly different IRF8 transcript abundance between these two groups.

## 3.0: Results

### 3.1: IRF8 abundance is elevated in monocyte-derived myeloid sarcoma

In order to investigate IRF8 expression in tumors of monocytic origin, we evaluated its abundance in myeloid sarcoma, an extramedullary malignancy of neoplastic immature monocytes and/or granulocytes (9). We performed immunohistochemical staining of 35 myeloid sarcoma patient samples and concomitantly stained for CD34, a hematopoietic stem cell marker, and myeloperoxidase (MPO), a myeloid marker. IRF8 was detectable in 24 of 35 (68.5%) of these cases, and CD34 was often undetectable in cases with the greatest degree of IRF8 positivity (Figure 1). Importantly, there were six cases for which IRF8 positivity was observed in the absence of CD34 or MPO expression, suggesting that IRF8 is able to identify myeloid sarcomas of primarily monocytic origin (Figure 2). In order to confirm monocytic origin of these blasts, double staining with lysozyme was performed on select MPO-negative specimens with sufficient remaining tissue. These showed blasts that were IRF8+ lysozyme+ as well as background scattered mature monocytes that were IRF8-lysozyme+ (Figure 3).

**Figure 1:**
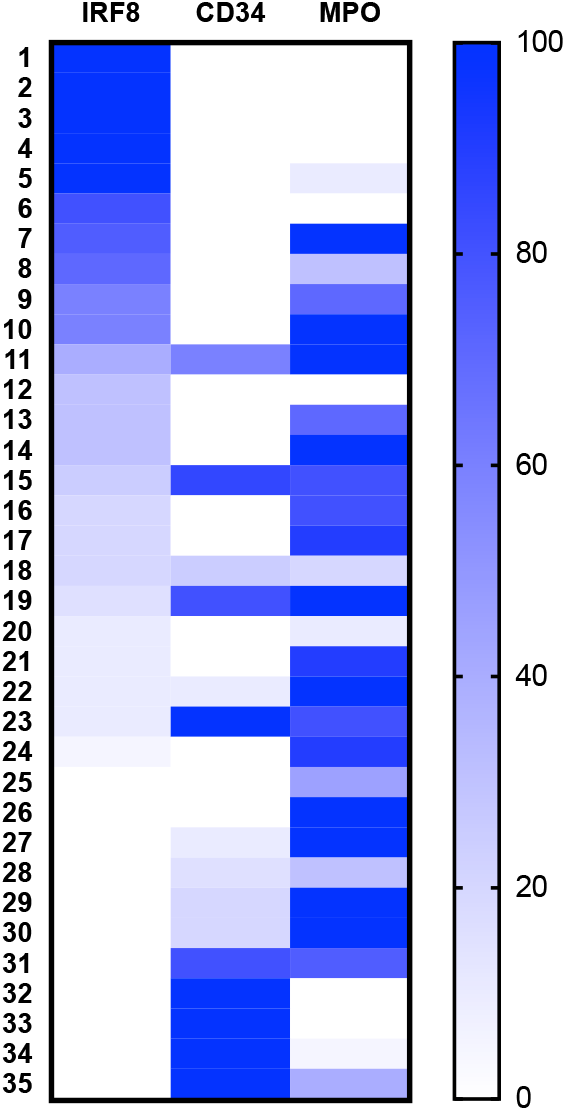
IRF8 expression in myeloid sarcoma. Heat map of myeloid sarcoma cases based on percent positivity of tumor cells for IRF8, CD34, or MPO by immunohistochemistry.

**Figure 2:**
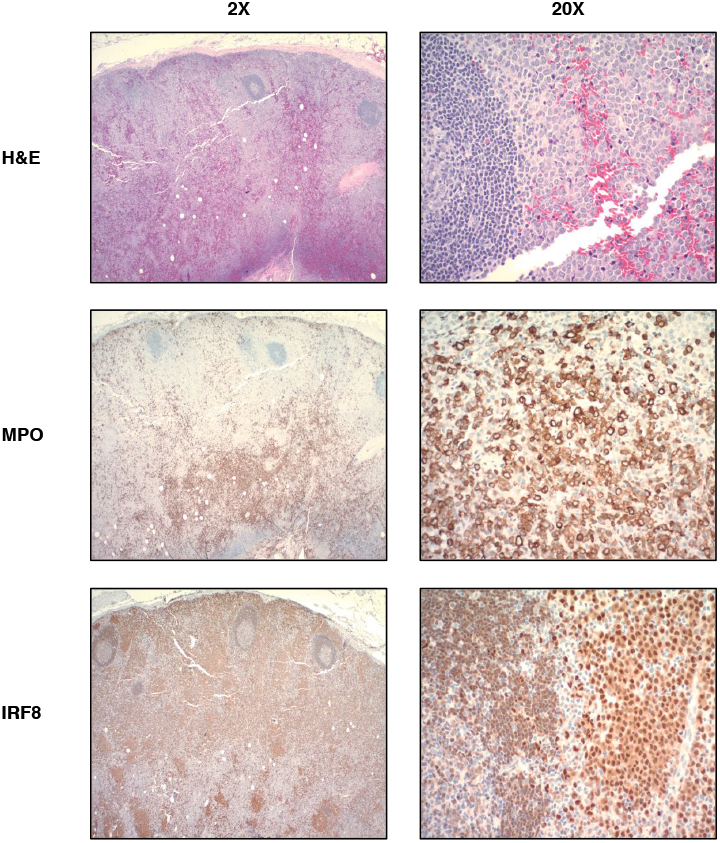
Representative case of myeloid sarcoma at 2X, 20X magnification. This tumor shows strong IRF8 expression in the monoblastic population of tumor cells and MPO staining in the granulocytic population (Case 8 in Figure 1). IRF8 expression is dimly observed in the mantle zone B cells adjacent to neoplastic cells.

**Figure 3:**
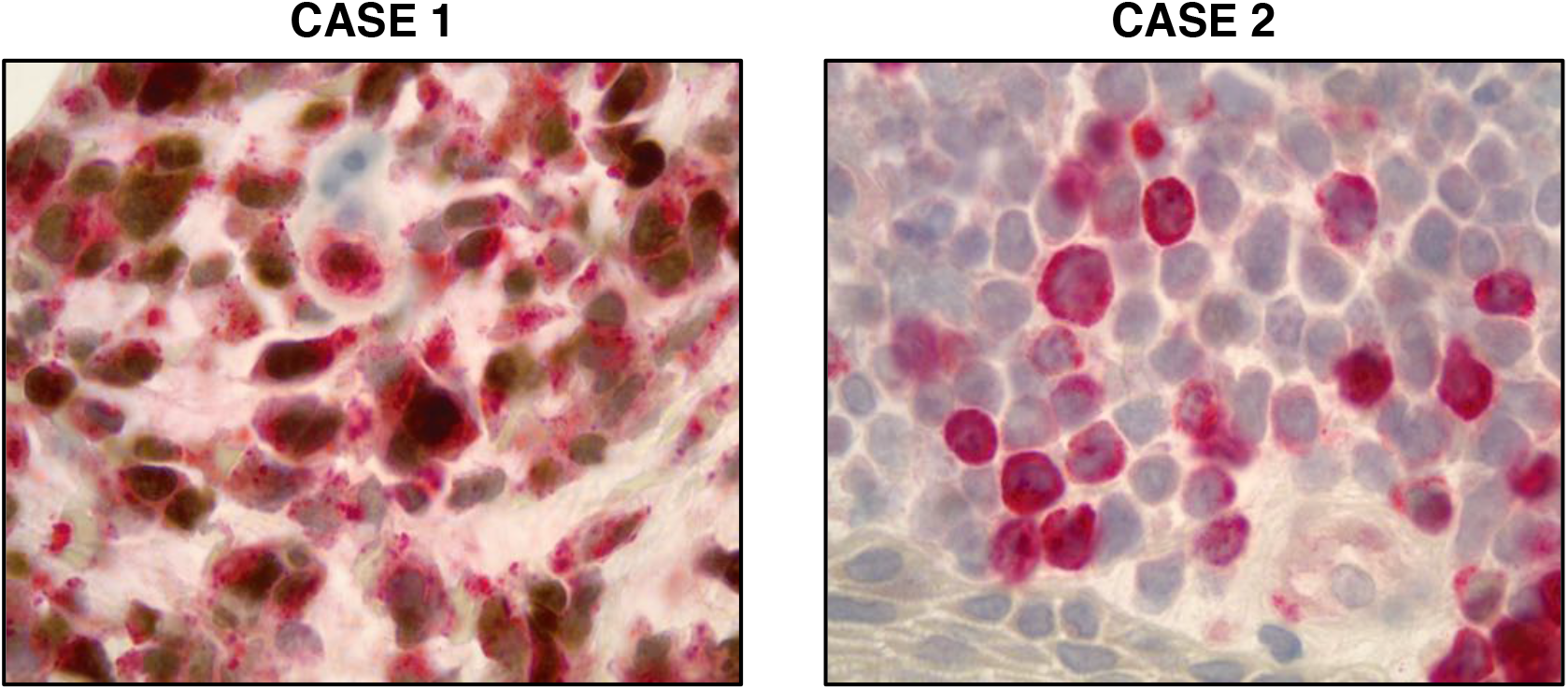
IRF8 and lysozyme staining in monocytic malignancies. Double stain with IRF8 (brown) and lysozyme (red) highlights double-positive monoblasts in a case of myeloid sarcoma (Case 1) and lysozyme single-positive monocytes in the skin (Case 2).

### 3.2: IRF8 is expressed in blastic plasmacytoid dendritic cell neoplasm

After observing IRF8 upregulation in myeloid sarcoma, we next investigated IRF8 abundance in blastic plasmacytoid dendritic cell neoplasm (BPDCN), a rare hematopoietic malignancy arising from immature plasmacytoid dendritic cells (17). Staining of 13 BPDCN cases demonstrated uniform IRF8 expression, with 13/13 (100%) samples showing positive IRF8 staining by immunohistochemistry. Importantly, there were multiple cases that stained negative for CD123, a marker commonly used in the diagnosis of BPDCN, but positive for IRF8 (Figure 4). This finding suggests that IRF8 holds promise as a marker for multiple difficult-to-diagnose hematologic malignancies of monocytic or dendritic cell origin.

**Figure 4:**
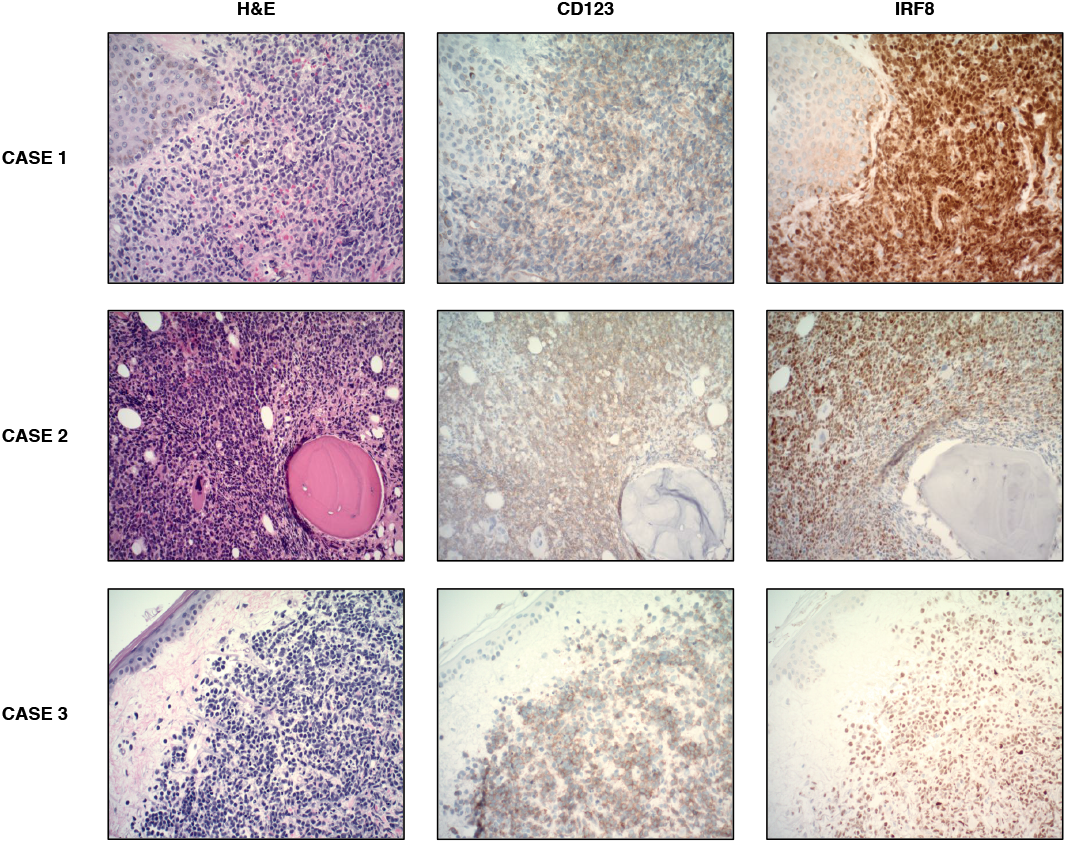
Expression patterns of BPDCN in three representative patients. Case 1 shows a mid-chest skin biopsy. Case 2 shows a bone marrow biopsy. Case 3 shows a left chest skin biopsy. All images taken at 200x magnification.

### 3.3: Assessment of IRF8 expression across different cancer types

We next sought to evaluate IRF8 expression across additional tumor types and normal tissues in order to validate that presence of IRF8 is specific to hematopoietic malignancies. Consequently, we performed IRF8 staining of a pan-cancer TMA comprising 177 patient samples of both normal and cancerous tissue across 11 tissue types and 25 cancer subtypes. IRF8 expression was negative in all normal tissue samples and all but one tumor sample, which was a diffuse large B-cell lymphoma of the testis (Table 1, Figure 5). We also sought to further evaluate IRF8 expression in breast cancer, as our small cohort of breast cancers in the initial pan-cancer TMA were devoid of triple-negative breast cancers (TNBCs). We additionally assessed IRF8 expression on a TMA of 97 TNBCs and observed that 2 (2.1%) of these tumors were IRF8-positive. The staining pattern for these cases was weak and showed subset reactivity (Figure 5). In many samples, we observed IRF8 staining of background lymphocytes that spared large, neoplastic cells.

**Table 1:**
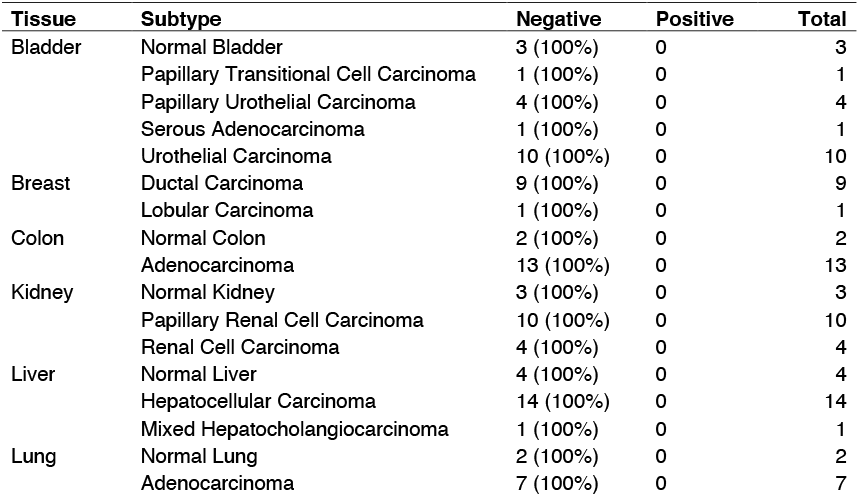
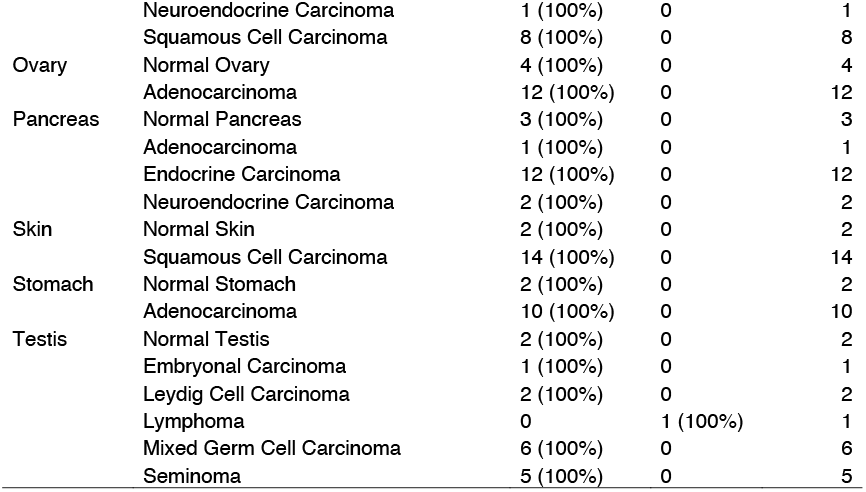
IRF8 expression in normal tissues and malignancies of different origins included in a pan-cancer TMA.

**Figure 5:**
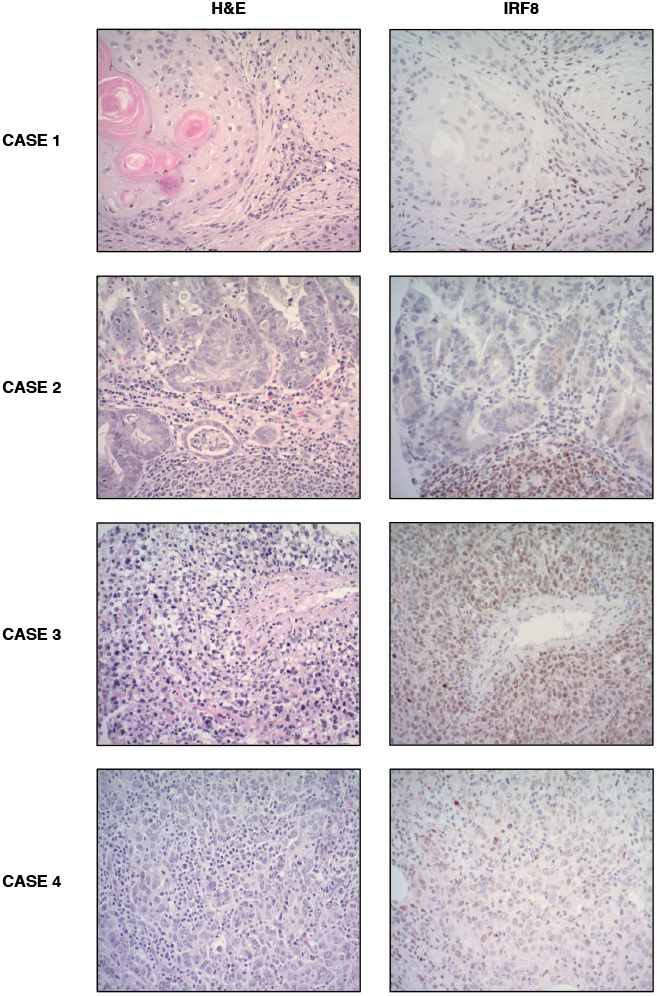
Representative cases of IRF8 immunohistochemistry from pan-cancer TMA. Case 1 shows a squamous cell skin carcinoma with IRF8 staining of infiltrating lymphocytes but not neoplastic cells. Case 2 shows a gastric adenocarcinoma with a similar staining pattern to Case 1. Case 3 shows a testicular lymphoma with IRF8 staining of large, polymorphic tumor cells. Case 4 shows a representative case of an IR8-positive TNBC.

We next analyzed publicly available expression data from The Cancer Genome Atlas (TCGA) to further assess IRF8 expression between normal and cancerous tissues for malignancies included in the TMA. In line with previous reports, IRF8 was significantly upregulated in acute myeloid leukemia and diffuse large B-cell lymphoma samples relative to normal tissue (7). However, IRF8 mRNA expression was also increased in colon adenocarcinoma, stomach adenocarcinoma, and testicular germ cell tumors (Figure 6). While these tumors were represented in our pan-cancer TMA, we did not observe a similar upregulation of IRF8 protein in the neoplastic cells, but rather in infiltrating lymphocytes (Table 1).

**Figure 6:**
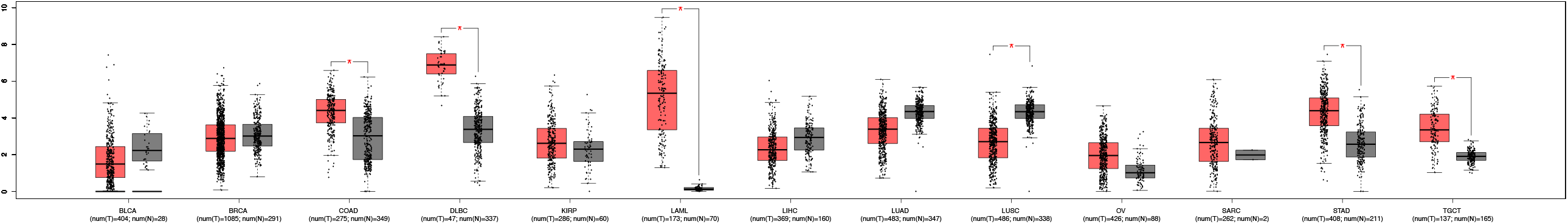
IRF8 expression in cancerous tissues (red) and normal tissues (gray). Box plots marked with a red asterisk indicate a statistically significant difference (p < .05) between tumor and normal tissue. BLCA – Bladder Urothelial Carcinoma, BRCA – Breast Invasive Carcinoma, COAD – Colon Adenocarcinoma, DLBC – Lymphoid Neoplasm Diffuse Large B-Cell Lymphoma, KIRP – Kidney Renal Papillary Cell Carcinoma, LAML – Acute Myeloid Leukemia, LIHC – Liver Hepatocellular Carcinoma, LUAD – Lung Adenocarcinoma, LUSC – Lung Squamous Cell Carcinoma, OV – Ovarian Serous Cystadenocarcinoma, SARC – Sarcoma, STAD – Stomach Adenocarcinoma, TGCT – Testicular Germ Cell Tumors.

We next performed IRF8 staining on sarcoma TMAs of subtypes that may be considered in the differential diagnosis of myeloid sarcoma or BPDCN. IRF8 expression was not observed in any of the 111 soft-tissue tumors belonging to 4 subtypes of sarcomas in these TMAs, which included desmoplastic small round cell tumor, Ewing sarcoma, synovial sarcoma, and undifferentiated pleomorphic sarcoma (Table 2). These results indicate that IRF8 expression is specifically upregulated in monocyte-derived soft-tissue tumors and can be utilized to identify myeloid sarcomas that lack CD34 or MPO expression. Taken together, these findings suggest that increased abundance of IRF8 has the potential to distinguish malignant cells across several cancer types and is most significantly associated with hematologic malignancies.

**Table 2:** IRF8 expression in different types of sarcoma included in sarcoma-specific TMAs.

| **Sarcoma Type** | **Negative** | **Positive** | **Total** |
| --- | --- | --- | --- |
| Desmoplastic small round cell tumor | 30 (100%) | 0 | 30 |
| Ewing sarcoma | 24 (100%) | 0 | 24 |
| Synovial sarcoma | 45 (100%) | 0 | 45 |
| Undifferentiated pleomorphic sarcoma | 12 (100%) | 0 | 12 |

## 4.0: Discussion

Malignancies derived from monoblasts are often difficult to diagnose, owing to their occasional morphologic overlap with other malignancies as well as a dearth of reliable immunohistochemical stains for these early hematopoietic progenitors (27). We recently identified IRF8 as a sensitive and specific monoblast marker in cases of acute monocytic leukemia (AMoLs) by demonstrating its strong correlation with aspirate blast count in AMoLs (8). IRF8 is a transcription factor that upregulates genes critical for the differentiation of monocytes, dendritic cells, and macrophages (2). Prior work has demonstrated that IRF8 expression peaks in monocyte and dendritic cell precursors, and thereafter remains high in mature dendritic cells but rapidly decreases in mature monocytes, suggesting it will potently stain immature monocytic/dendritic cell populations (3, 28). Herein, we have demonstrated that IRF8 is expressed in multiple hematologic malignancies of monoblastic/dendritic cell origin and holds promise as a biomarker for these difficult-to-diagnose neoplasms.

Given our recent work demonstrating IRF8 as a reliable monoblast marker, we sought to evaluate the utility of IRF8 in staining monocyte-derived hematopoietic cancers for which clinical diagnosis is challenging and helpful tissue markers are lacking. We first stained for IRF8 in samples of myeloid sarcoma, an extramedullary tumor of mixed immature granulocyte and monocyte origin (15). Myeloid sarcoma can rarely occur as an isolated disease but is most commonly observed as a manifestation of acute myeloid leukemia, being reported in roughly 2.5-9.1% of AML patients (11). It is often difficult for pathologists to diagnose, particularly in the absence of a previous AML diagnosis; prior studies have estimated that between 25-47% of myeloid sarcoma cases are misdiagnosed, largely due to inadequate immunophenotyping of the lesion (12-14). Consequently, we posited that IRF8 could be utilized to stain myeloid sarcoma cases with prominent monoblast populations. IRF8 was able to stain a significant proportion of myeloid sarcoma cases, including those negative for CD34 and MPO; these cases that had previous/concurrent marrows did indeed demonstrate monocytic differentiation by aspirate morphology with or without flow immunophenotypic support. Additionally, IRF8 did not stain any other soft-tissue tumors on sarcoma TMAs representing some of the differential diagnoses of myeloid sarcoma, including Ewing sarcoma, undifferentiated pleomorphic sarcoma, desmoplastic small round cell tumor and synovial sarcoma (29). Thus, IRF8 can be utilized as a biomarker for myeloid sarcoma and has the potential to improve diagnostic accuracy of this less common hematologic malignancy, particularly for cases which are primarily of monocytic origin.

We next evaluated the utility of IRF8 in staining malignant cells in blastic plasmacytoid dendritic cell neoplasm (BPDCN). BPDCN is an aggressive, rare tumor derived from precursors of plasmacytoid dendritic cells that often presents asymptomatically or with multiple bruise-like cutaneous lesions before progressing to involve extracutaneous sites (20). In roughly 66% of patients, BPDCN presents as stage IV disease at time of diagnosis, and median survival is estimated to be between 12 and 14 months (22). BPDCN diagnosis is made on the basis of clinical features, morphological findings, and immunophenotype. Diagnosis is challenging, and BPDCN is often difficult to distinguish from soft-tissue tumors, AML, and T-cell leukemias/lymphomas. This is due to both an overlap in immunophenotyping as well as the common lymphoid-like morphology of BPDCN blasts that leads to a false suspicion of lymphoma (30). Given the dendritic cell lineage of BPDCN blasts and the diagnostic difficulties associated with the disease, we hypothesized that IRF8 could be a useful tool in staining neoplastic BPDCN cells. All 13 samples we obtained of this highly rare tumor were strongly positive for IRF8 staining. Additionally, several of these IRF8-positive cases were negative for CD123, a marker commonly used for BPDCN, suggesting that IRF8 may be a useful marker to add to the diagnostic panel for this aggressive hematopoietic malignancy (31).

Following the identification of positive IRF8 staining in myeloid sarcoma and BPDCN, we sought to determine if its expression is specific to cancers of hematologic origin. We showed that IRF8 did not stain any solid tumors or normal tissue on a pan-cancer TMA comprising 25 cancer subtypes. However, global expression data from The Cancer Genome Atlas (TCGA) suggested that IRF8 is transcriptionally upregulated in several carcinomas relative to normal tissue, particularly colon adenocarcinoma, stomach adenocarcinoma, and testicular germ cell tumors, all of which were represented in our TMA. In contrast to this TCGA data, prior reports have indicated that IRF8 expression is preferentially decreased in both colon and gastric cancer samples relative to noncancerous tissue, through mechanisms such as increased promoter methylation (32, 33). This discrepancy highlights the discordance that can be observed between TCGA data and primary patient samples. Moreover, while breast cancers represented in the pan-cancer TMA were uniformly IRF8-negative, we performed additional staining on a TMA specific for triple-negative breast cancers (TNBCs) due to a recent study documenting increased IRF8 expression in estrogen receptor-negative breast cancers (34). However, only rare cases (2.1%) of the TNBCs in this TMA stained weakly positive for IRF8. This observation is potentially due to the fact that these investigators found IRF8 expression to be increased specifically in early-stage ER-negative cancers, which may not have been as well represented on the TMA as advanced disease. Importantly, and in line with recent work, IRF8 upregulation was most significantly associated with AML and diffuse large b-cell lymphoma in the TCGA dataset, further indicating that IRF8 likely has greatest utility as a marker of hematopoietic neoplasms (7).

In this study, we demonstrate that IRF8 can be used to identify monocyte and dendritic cell-derived malignancies that are prone to misdiagnosis due to a scarcity of specific markers. Our study is limited by the relatively few cases of BPDCN. In addition, since B-cell lymphomas are also positive for IRF8, the marker does not help to differentiate among hematopoietic tumors. Further validation of these findings will be critical in order to integrate IRF8 staining into pathologists’ workflow for diagnosis of these diseases. Additionally, future studies should build upon this work by determining if differential IRF8 expression correlates with poor prognosis or response to therapy in the diseases it stains. While this will likely prove challenging due to the rarity of both myeloid sarcoma and BPDCN, identifying additional prognostic markers for these diseases is crucial, particularly given the lack of data regarding survival differences between granulocyte-predominant and monocyte-predominant myeloid sarcoma (35). IRF8 thus may hold promise not only as a marker for diagnosis but also as a prognostic indicator that guides treatment intervention for these rare hematopoietic neoplasms.

## Data Availability

All data produced in the present work are contained in the manuscript.

## Acknowledgements

We acknowledge Amos Brooks and Lori Charette from Yale Pathology Tissue Services for their invaluable experience and aid with immunohistochemical studies as well as tissue procurement and processing.

## Ethics Approval and Consent to Participate

The study was performed in accordance with the Declaration of Helsinki. The study was approved by an internal review board (local ethics committee HIC protocol #2000023891).

## Author Contributions

D.C.M., S.G.K., and M.L.X. contributed equally in conception and design. D.C.M., M.L.X, and G.P. carried out data analysis. W.W. contributed additional cases. D.C.M. wrote the manuscript with significant contributions from M.L.X. and S.G.K. All authors edited the manuscript.

## Data Availability

The tissue sections analyzed during this study are available from the corresponding author upon request. TCGA data analyzed during this study are publicly available using the <u>GEPIA online resource</u>.

